# International consensus on fluoride programmes for Early Childhood Caries prevention in early education settings

**DOI:** 10.1101/2024.03.05.24303843

**Authors:** Lamis Abuhaloob, David I Conay, Alex Blokland, Al Ross

## Abstract

**Background:** The World Health Organisation has set out a clear priority for the implementation of interventions to reduce the burden of Early Childhood Caries (ECC), a global public health problem affecting over 500 million children around the world and having a substantial impact on child well-being and health system expenditure.

The aim of this study was to assess and develop international expert consensus on the evidence for fluoride-based interventions in early-year education settings (kindergartens/nursery and primary schools) for reducing ECC and to synthesise clear programme-level recommendations with regard to ECC prevention in this setting.

**Methods:** A systematic overview of systematic reviews, trials, and observational studies was performed to identify and critically appraise the available evidence on the effectiveness and cost-effectiveness of fluoride-based interventions in early-years education settings to prevent ECC.

This was followed by a three-stage modified Delphi panel study (n= 21) consisting of: round 1, an online survey to gather opinions on safety, effectiveness and feasibility of interventions; round 2, an iterative survey to consider collated group opinion and gather feedback on review findings; and finally, an online workshop with presentations and facilitated in-depth, recorded group discussions.

**Results:** There was high-quality evidence and consensus on delivering supervised toothbrushing in kindergartens (nurseries) and primary/elementary schools; this is safe and cost-effective, shows greater benefit to more disadvantaged children, helps child social development, and is feasible in high and low/middle-income countries. There was more moderate support for the effectiveness and cost-effectiveness of fluoride varnish application in this setting (especially where supervised toothbrushing with fluoride is in place). It was agreed that policy makers should prioritise at-risk groups where resources are limited, and that systemic fluoride interventions (Fluoride tablets, drops, milk, and salt) in this setting are no longer a priority.

**Conclusion:** Supervised toothbrushing with fluoridated toothpaste is the most effective, cost-effective, feasible and safest mechanism for children in early education settings. Universal coverage is preferred but where resources are limited targeting based on need is indicated. Panel consensus is that it remains appropriate in water fluoridated areas and is largely feasible in low/middle-income countries.

## Introduction

The World Health Organisation (WHO) has set out a clear priority for the implementation of programmatic interventions to reduce the burden of Early Childhood Caries (ECC), a non-communicable disease and global public health problem affecting children around the world [1]. Recent reports indicate that 514 million children worldwide (43% of the population) have dental caries in their deciduous teeth, with three -quarters of those living with untreated decay living in low/middle-income countries [2].

In 2019 the WHO published an implementation manual for ending ECC [3] that outlined a number of important principles for preventive programmes, including involvement of primary care teams and recognition of the role of school-based programmes. Key fluoride-based interventions outlined in the manual which can be delivered in education settings are: routine brushing of infants’ and children’s teeth with fluoride toothpaste [4]; fluoridated salt or milk programmes [5–8]; and regular application of 5% sodium fluoride varnish [9, 10].

Fluoride varnish is currently recommended in clinical guidance as a safe, topical treatment for caries prevention in children [11] which is supported by high quality reviews [9]. However, some new emerging evidence needs to be considered. A recent systematic review [12] concluded that “there is modest and uncertain evidence for fluoride varnish reducing the risk of developing dentine caries in pre-school children”. Similarly, a recent double-blind, two-arm randomised controlled trial embedded within the Childsmile programme (Protecting Teeth at Three; PT@3) found a modest non-significant reduction in the worsening of d3mft in the nursery Fluoride varnish group compared to Treatment as Usual (universal supervised toothbrushing) in the nursery setting for children at high risk, concluding that it was neither effective or cost-effective [13, 14].

Studies of effectiveness generally show improvement in the dental health of young children associated with exposure to toothbrushing with fluoridate toothpaste in the education setting. Cost analysis to inform the development of national nursery toothbrushing programmes in Scotland showed a cost saving of approximately GBP6.0 million over eight years compared with health service costs associated with dental treatments [13, 14].

Evidence for systemic fluoride in education settings largely predates the use of topical fluoride. The effectiveness of fluoride supplements (tablets, drops, lozenges) on deciduous teeth (nursery or early primary school children) appears unclear (there is some reported reduction in caries increment for older children with permanent teeth) and there is limited information on adverse effects. Similarly, evidence on fluoridated salt (typically containing 250ppm fluoride) is substantially over 20 years old, but supportive in children at primary/elementary school age and above. There is some evidence for effectiveness of fluoridated milk (versus non-fluoridated milk) in longitudinal cohorts but concerns about risk of bias and applicability to different populations.

Thus, prevention of dental caries in young children is an agreed priority for policy-makers due to the high disease levels and potential savings from associated expensive treatment costs, and delivery of fluoride in education settings can play a role. There are questions remaining about the best intervention(s) in terms of safety, efficacy and feasibility, and therefore there is need for appraisal of the relative merits and demerits of toothbrushing, fluoride varnish, and other fluoride-based programmes for children in education settings as part of population oral health improvement policies and programmes.

### Aims

This study aimed to assess and develop an international expert consensus on fluoride-based interventions in early years educational settings for reducing ECC in young children.

The objectives were:

- Collate and evaluate up-to-date scientific evidence and clinical guidance on fluoride varnish and toothbrushing interventions for the prevention of ECC,
- Determine expert consensus on fluoride-based interventions in early years educational settings,
- Synthesise and summarise findings in a concise and accessible form to inform implementation efforts.

## Materials and Methods

The study was a mixed-methods project with a systematic overview and appraisal of evidence being fed into iterative consensus-building ‘rounds with a mixture of quantitative and qualitative procedures.

### Systematic overview

The overview was registered in the International prospective register of systematic reviews [15]. A systematic literature search syntax was developed for MEDLINE and adapted for the following databases: EMBASE, Web of Science Core Collection, and Cumulative Index to Nursing and Allied Health Literature (CINAHL). Filters based on the recommended SIGN filters were used to target the search at interventions (trial and evaluation papers).

### Inclusion/ exclusion criteria

- All peer-reviewed study designs were included (i.e. systematic reviews, Randomised Controlled Trials, observational designs)
- No restrictions on language or publication dates were placed
- Studies were restricted to those delivered directly in early education settings (nursery/kindergarten or primary/elementary school).
- Only studies of fluoride-based interventions (e.g. fluoride toothpaste, fluoride gel or foam, fluoride varnish, fluoride rinsing, fluoride tablets, and supplements in milk) were included
- No restrictions were made on comparators or reported outcomes

We excluded study protocols, systematic review protocols, non-peer reviewed reports and conference abstracts for which no full text was available. We also excluded minimally invasive techniques such as Silver Diamine Fluoride (SDF) and Atraumatic Restorative Treatment (ART).

### Study appraisal and data extraction

The cohort of identified papers (Fig 1) was managed in Covidence Systematic Review Software [16]. Title and abstract screening and full text screening was carried out by at least two independent reviewers. Any ambiguity/disagreements were discussed between all three reviewers to reach an agreement. For excluded papers with reasons for exclusion see S1Table.

**Fig 1.**
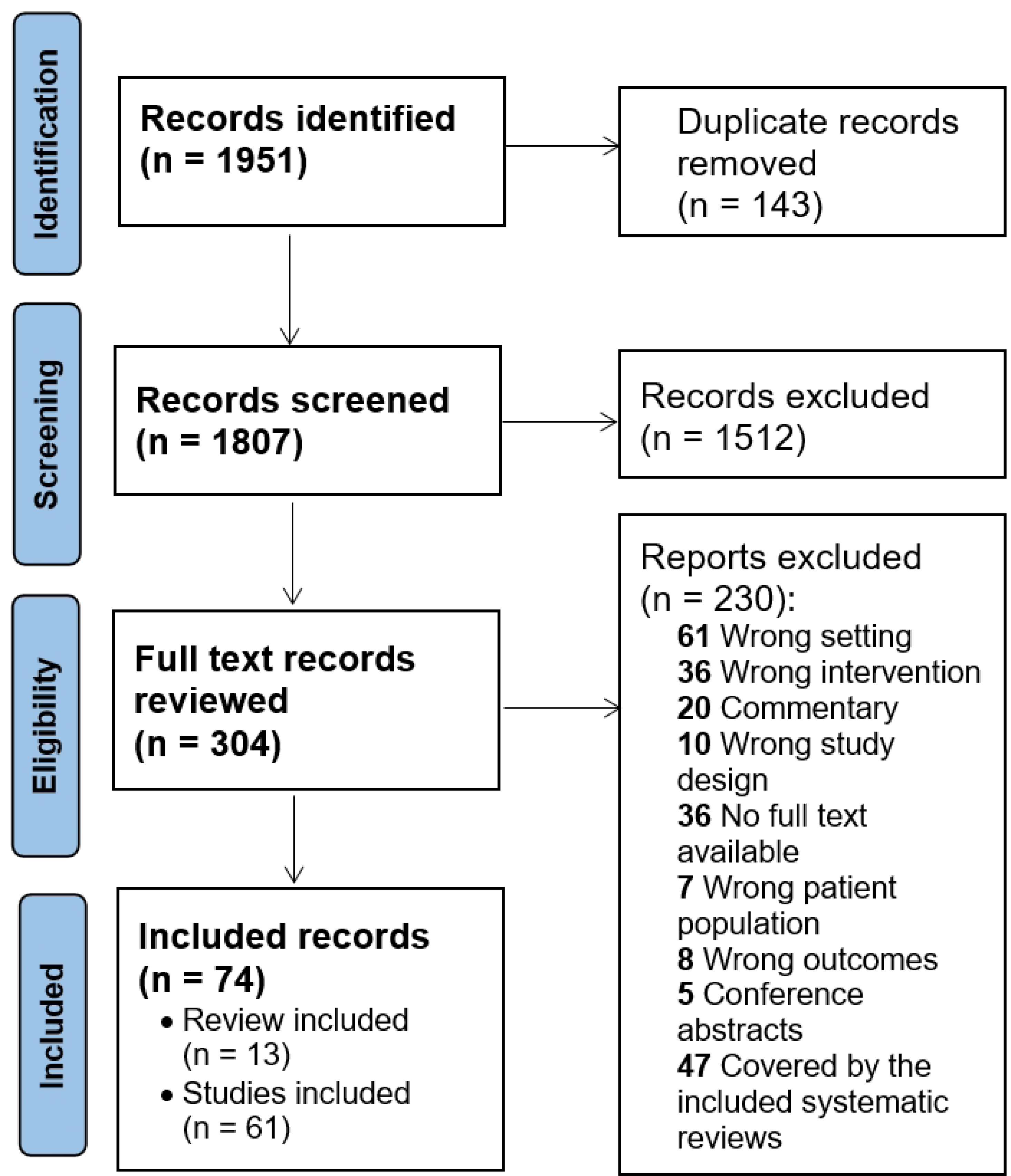
PRISMA flow diagram for the systematic overview of fluoride interventions in early education settings.

Quality appraisal and assessment of the risk of bias was again undertaken independently by at least two reviewers, after which disagreements were discussed, using appropriate tools for systematic reviews (AMSTAR and ROBIS) and primary studies (CASP) [17–19].

Data extraction included capturing: study design; study setting; target population; sample size; brief description of the intervention; comparators / treatment as usual; clinical outcomes (efficacy, safety) and/or cost/economic outcomes.

Because of the high heterogeneity (setting/population, intervention/control, duration, outcomes) random effects meta-analysis could not be performed and data synthesis is in narrative form. Results which were fed into the Delphi rounds were extracted from high and medium quality papers – starting with Systematic Reviews, supplemented by further evidence from separate primary studies not included in reviews. After quality, recency of data was prioritised. Lower quality evidence is summarised briefly.

### The Modified Delphi consensus-building exercise

The Modified Delphi exercise followed conventional steps for remote/online consensus building [20]. The methodology is a validated systematic method to measure and develop consensus when empirical evidence is emerging, lacking, limited, or contradictory [21].

There were three rounds to the Modified Delphi exercise. First, the panel were sent an initial survey to gather ‘baseline’ opinions on safety, efficacy and priorities for implementation in both high and low/middle-income settings. For ‘round 2’, survey proposition statements from the systematic overview were fed in for opinion as well as collated priority data, and selected qualitative opinions, from round 1. Survey results helped set the questions/scope for a final online meeting of the group (workshop; around 150 minutes). This involved presentations from the research team with questions/answers and then three parallel ‘break out’ facilitated discussions.

### Procedures

Participants were purposively selected due to their expertise and their ability to further the aims of the work. They were targeted through existing connections: senior researchers and clinical academics including the Cochrane Oral Health Group; Dental Public Health leads; senior Government and policy/programme leads in childhood caries prevention.

Data gathering for the Delphi consensus exercise was through MS Forms, hosted on the University of Glasgow’s secure OneDrive, with access restricted to the named project team. Workshop was facilitated through MS Teams. The meeting was password protected. Teams allows recording directly to the University server (OneDrive) and storage of sessions without recourse to third-party software. All data was stored on a secure university drive on a password protected computer, in accordance with the University of Glasgow’s data security protocol which is in full compliance with the General Data Protection Regulation (GDPR 2018).

Workshop consent was secured by email in advance. The workshop discussion was recorded and transcribed. Transcripts were disidentified with quotations attributed to unique participant ID numbers only.

Surveys responses were analysed by way of descriptive statistics in IBM SPSS Statistics 26 software and presented into stacked bar chart. Workshop’s discussions were transcribed, and were analysed using thematic analysis based on the Consolidated Framework for Implementation Research [22] facilitated by QSR NVivo 12.0 data analysis software.

Study ethical approval was granted by the College of Medical, Veterinary and Life Sciences Ethics Committee, University of Glasgow (Project No: 200210053).

## Results

Fig 1 shows 1951 initial papers were identified from the databases; 304 (Fig 1) were appraised at full text screening and a final 74 papers were included for quality appraisal and data extraction: 13 systematic reviews including 3 economic reviews; 23 randomised controlled trials; 29 observational studies; and 9 economic studies.

Extracted data from all primary studies (design, population, setting, sample, outcomes and key findings) and reviews (type, quality appraisal, interventions, outcomes, key findings, education setting findings where distinct) in the overview are shown in S2 and S3 Tables.

Results of the overview and Delphi exercise are summarised below by intervention, with a focus on high and moderate quality papers. Twenty-one experts responded to the first round Delphi survey and fifteen to the second round (71%). Qualitative feedback from the panel for all interventions was then summarised thematically.

### Comparing interventions

The panel were asked, based on their opinions on effectiveness and safety, to rank the interventions in terms of priority as part of efforts to reduce early childhood caries, where 1 = highest priority and 7 = lowest priority. Results are shown in Fig 2.

**Fig 2.**
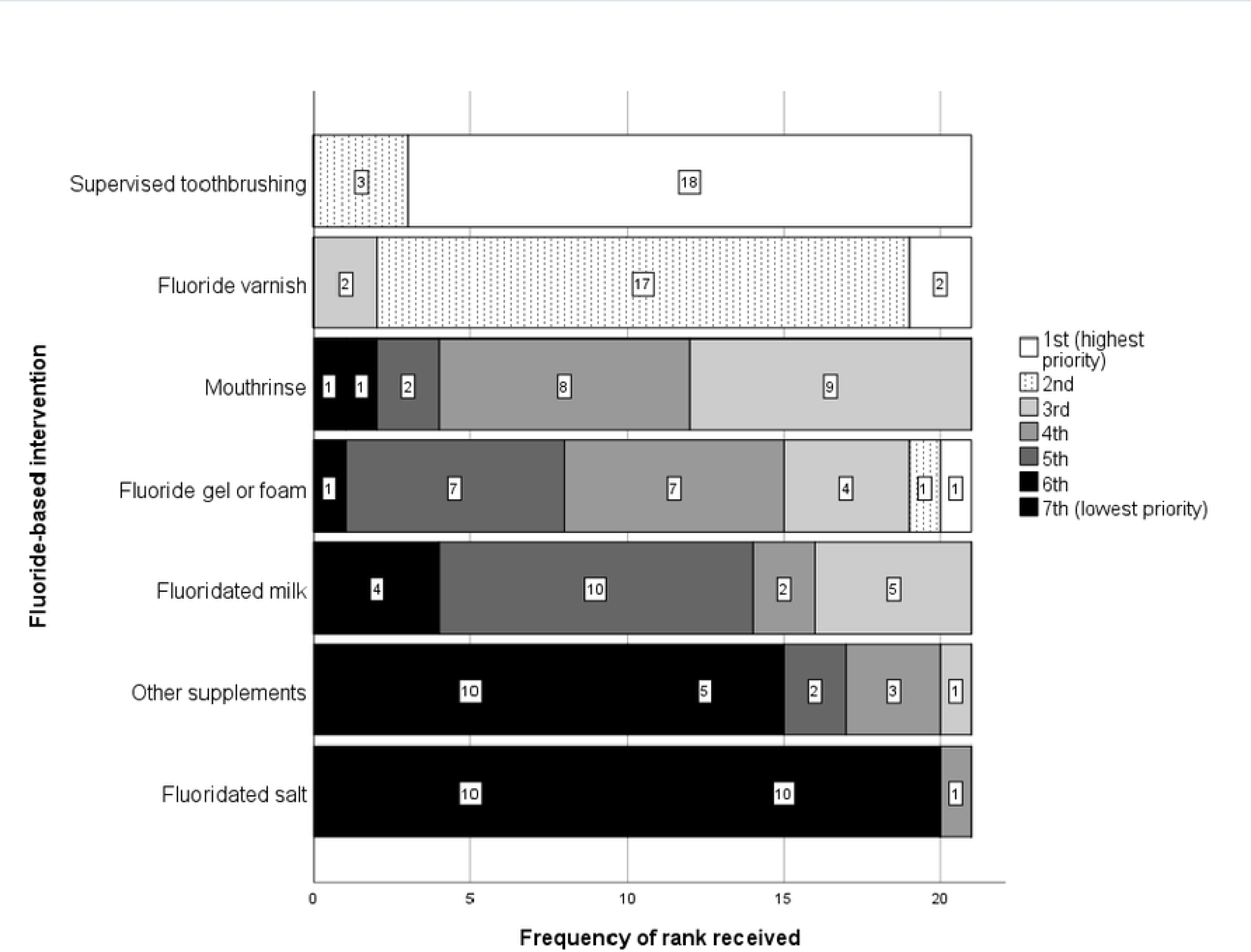
Expert panel ranking of fluoride interventions in nurseries/schools based on effectiveness and safety (n= 21)

The panel were then asked, taking into account cost and other potential barriers, to rank the interventions in terms of their feasibility of implementation in high income, and low/middle income countries separately, where 1 = highest priority and 7 = lowest priority.

Results for high income countries are shown in Fig 3. The results for low/middle-income countries (Fig4) were similar, with changes restricted to: fluoride gel or foam down from rank 4 to 5; fluoridated milk up from 5 to 4.

**Fig 3.**
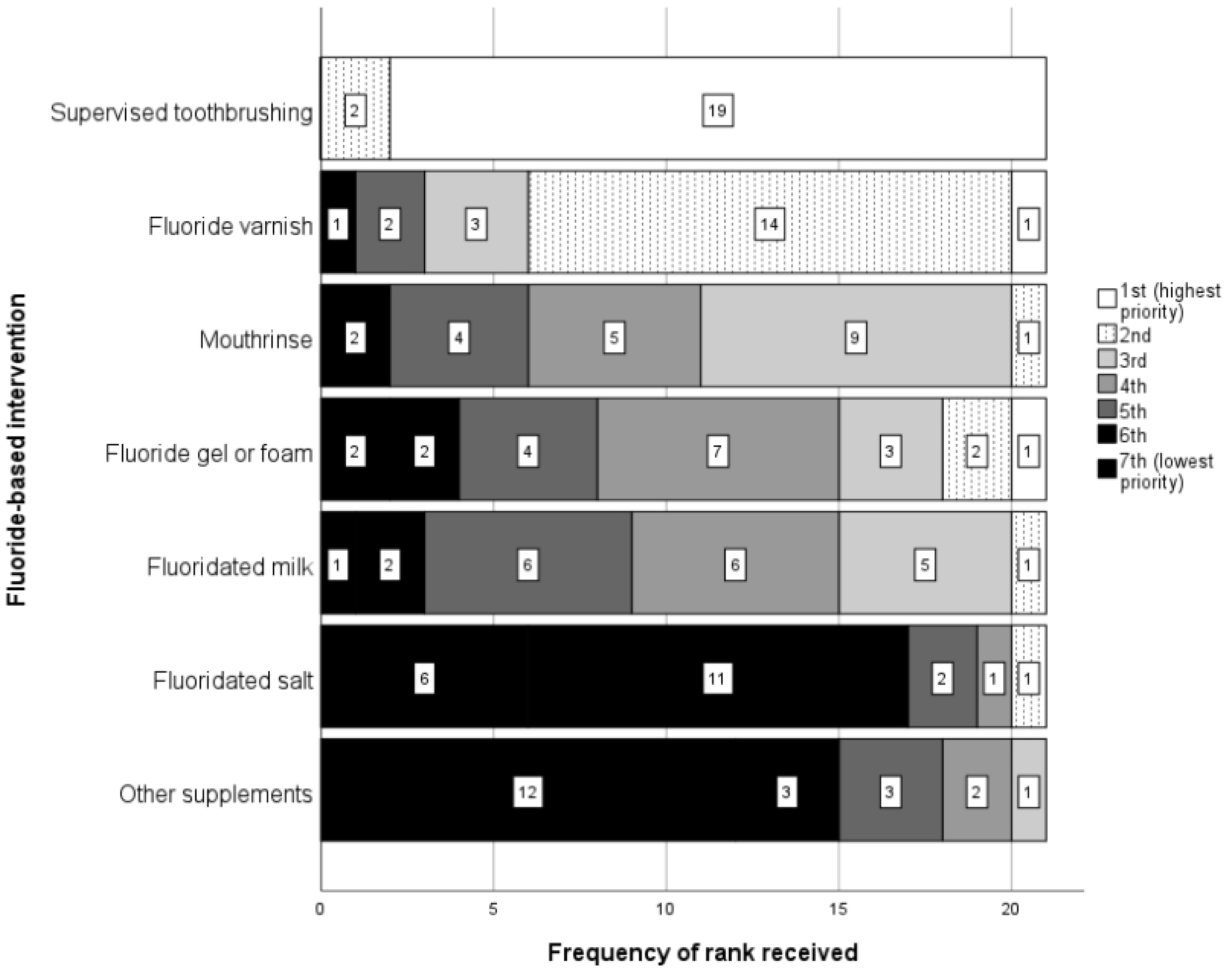
Expert panel ranking of fluoride interventions in nurseries/schools based on feasibility of implementation in high income countries (n= 21)

**Fig 4.**
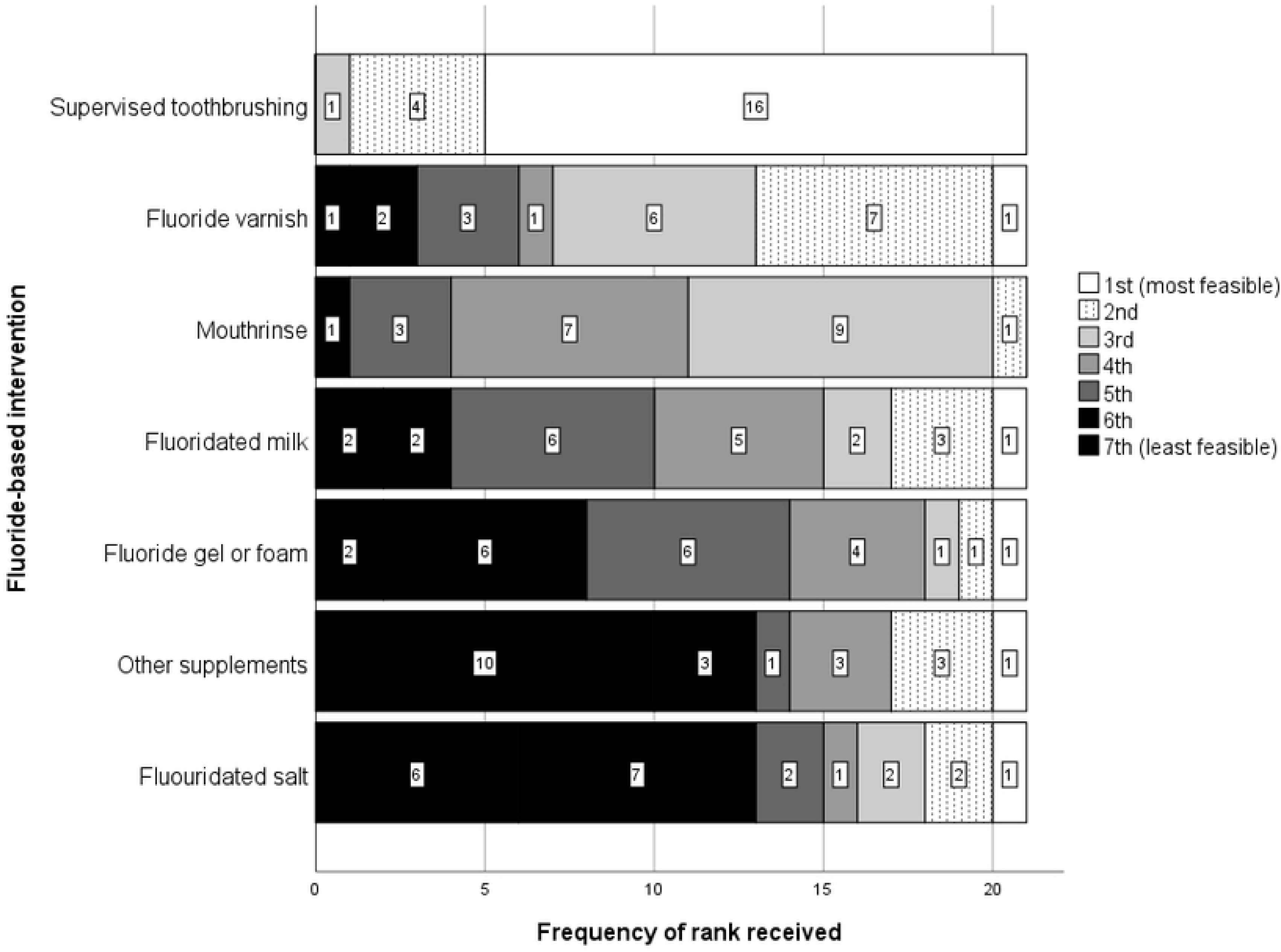
Expert panel ranking of fluoride interventions in nurseries/schools based on feasibility of implementation in low/middle income countries (n= 21)

### Supervised daily toothbrushing with fluoride toothpaste

It can be seen from Figs 2 and 3 that daily supervised toothbrushing with fluoride toothpaste was clearly the top-ranked intervention, with 18/21 (86%) ranking it as the top priority in terms of effectiveness and safety and 19/21 (90%) ranking it as the most feasible in high income countries. In low/middle-income countries 16 (76%) ranked it first and 4 (19%) ranked it second. No other intervention received more than one vote for top rank.

This collated opinion was closely aligned with strong supportive evidence from the systematic overview. As can be seen from S2 Table, studies showed a significant reduction (33-40%) in dental caries increment with daily supervised toothbrushing using 1000 to 1450 ppm Fluoride toothpaste in children aged 2-12 years old in nurseries and primary schools [4, 23, 24].

A recent high-quality economic study of a national supervised toothbrushing programme in nursery schools in Scotland found that after eighth year of the toothbrushing implementation, the expected savings (£4,731,097) were more than two and a half times the costs of the programme implementation (£1,762,621 per year).

Furthermore, a population standardised analysis by deprivation groups showed that the largest decrease in modelled costs was for the most deprived cohort of children [14]. There is limited evidence for supervision *per se* (against unsupervised brushing, see thematic findings). Additionally, an indicative qualitative survey response was *‘I still hold daily toothbrushing with fluoridated toothpaste will confer more consistent protection against dental caries, when compared to periodic delivery of fluoride through varnish’*.

In round 2, 13/15 respondents (87%) agreed or strongly agreed with the proposition that *‘toothbrushing with fluoride toothpaste should be universal for all children in early years education’.* However, in terms of targeting, 11/15 (73%) agreed or strongly agreed that *‘if resource is limited, [it is] fair enough to only offer [toothbrushing] in areas of relative deprivation’*.

### Fluoride varnish application

Fluoride varnish application (usually 2-4 times yearly, see S2 Table) was consistently the second-ranked intervention, with two top ranks for effectiveness/safety and 19/21 (90%) ranking it first or second priority. This intervention received one first and 14 (67%) second rankings for feasibility in high-income countries. In low/middle-income countries, fluoride varnish was still clearly ranked second in terms of feasibility to daily supervised toothbrushing, but the percentage of first or second rank fell from 15/21 (71%) to 8/21 (38%).

In round 2, just one person reported that the overview summary matched their own understanding of the evidence for fluoride varnish in nurseries and schools ‘to a very large extent’. Eleven out of 15 (73%) did agree it matched ‘to a large extent’ whilst two were unsure and one said it matched ‘to very little extent’. This compared to six people (40%) who felt the toothbrushing overview matched their own understanding ‘to a very large extent’.

The overview demonstrated that fluoride varnish delivered in nurseries or primary schools typically focuses on two-to-four applications a year of sodium fluoride varnish containing around 22,600 ppm fluoride. One RCT reported a 49% reduction in dental caries (P< 0.001) in high-risk pre-school children receiving fluoride varnish four times in Kosovo [25]. Two other high-quality trials reported a modest non-significant reduction in the worsening of d3mft in a low socioeconomic nursery setting in Scotland [13]; and no significant caries-preventive effect in a primary school setting within a high-risk community in South Africa [26].

A more recent fluoride varnish review - not restricted to education settings, which was rated as moderate quality, showed a modest and uncertain anti-caries effect in pre-schoolers and called for more cost-effectiveness analyses [27].

The highest quality review shows mixed (positive, negative and inconclusive) results in the education setting where there is a paucity of high-quality randomised trials [9]. In Survey 2, 93% of respondents declared that this evidence matched their understanding of the evidence about fluoride varnish in early education settings. Three respondents illustrated that they don’t have a full understanding of observational studies and were unaware of the negative results on cost-effectiveness.

Our review was feeding in a question about fluoride varnish to the panel. As one qualitative survey response had it, *‘you have told me it is not as strongly supported by evidence as I previously thought’.* When asked whether they agreed that “There is questionable value in continuing fluoride varnish in the early education setting where children are undergoing supervised toothbrushing with fluoride”, four strongly agreed, three agreed, seven (47%) were ‘not sure’ and one person disagreed. This suggest this is an area where consensus is somewhat lacking and in need of further consideration.

### Other fluoride-based interventions

There was broad consensus that other fluoride-based interventions are less of a priority in terms of effectiveness/safety and feasibility (Figs 2 and 3). The data extracted during the review (S2 and S3 Tables) show, for example, that there is limited information on the adverse effects of fluoride mouth rinse application in educational settings to reduce ECC [28, 29]. The reported caries preventive benefit was larger among children in high-risk schools [30] and in areas of non-fluoridated water [31]. High-quality evidence showed that cost-effectiveness is increased where teachers supervise such programmes [32]. However, a highly rated Cochrane review reported that the size of the preventive effect of such interventions in school settings is uncertain [33].

Fluoride gel (typically applied quarterly, containing up to 12,300ppm fluoride) effectiveness in reducing dental caries in permanent dentition was supported with moderate quality evidence; there is low-quality evidence for caries prevention via fluoride gel in primary dentition and there is again limited information on its adverse effects [34].

Similarly, it is difficult to draw definitive conclusions about the benefits of milk fluoridation (typically containing 250ppm fluoride) in education settings. [35]. A parallel arm longitudinal cohort study in Bulgaria reported a 61% reduction in caries permanent teeth from a community milk fluoridation programme [8]; concerns remain regarding the generalisability of these findings.

Delphi results confirmed the view that fluoride supplement evidence substantially predates the use of topical fluoride. The effectiveness of fluoride supplements on deciduous teeth (nursery or early primary school children) appears unclear and there is limited information on adverse effects [36]. Evidence on fluoridated salt (typically containing 250ppm fluoride) is substantially over 20 years old but supportive in slightly older children (primary school age and above aged 10-12 years old) [37]. In a more recent study in Gambia - a country [38] with low levels of fluoride in drinking water (0.1 mg F – /L) and a high caries burden, it was found that the use of fluoridated salt to prepare school meals resulted in 66.3% caries-preventive effect in preschool children [39]. Generally, there is unclear evidence on the effectiveness of administration of combined interventions in education settings versus single interventions.

In the survey, 100% of respondents agreed or strongly agreed that salt as a way of delivering fluoride should be questioned because it is a risk factor for hypertension. they illustrated that it is ‘*indicated [as a risk factor] in other non-communicable diseases and […] targeted for reduction in many foods*’. Ten out of 15 (67%) experts agreed or strongly agreed that there is insufficient high-quality evidence on mouth rinse to recommend it.

Finally, more experts (82%) agreed that fluoride-based interventions should be implemented over and above dental health education for children than those (58%) who agreed that fluoride-based interventions should be implemented over and above community water fluoridation. They collectively (87%) highlighted the vital value of environmental sustainability and cost-effectiveness data to inform policies on fluoride-based interventions in education settings. In addition, they agreed that fluoride varnish application in education settings would effectively tackle dental caries in high deprivation areas (80%), The majority of the experts (86%) recommend universal application of toothbrushing intervention in early education settings, irrespective of high/middle/low-income country status and irrespective of presence/absence of community water fluoride.

### Workshop discussions

Finally, Table 1 shows illustrations of Delphi workshop discussion groups, arranged by cross-cutting themes.

**Table 1.**
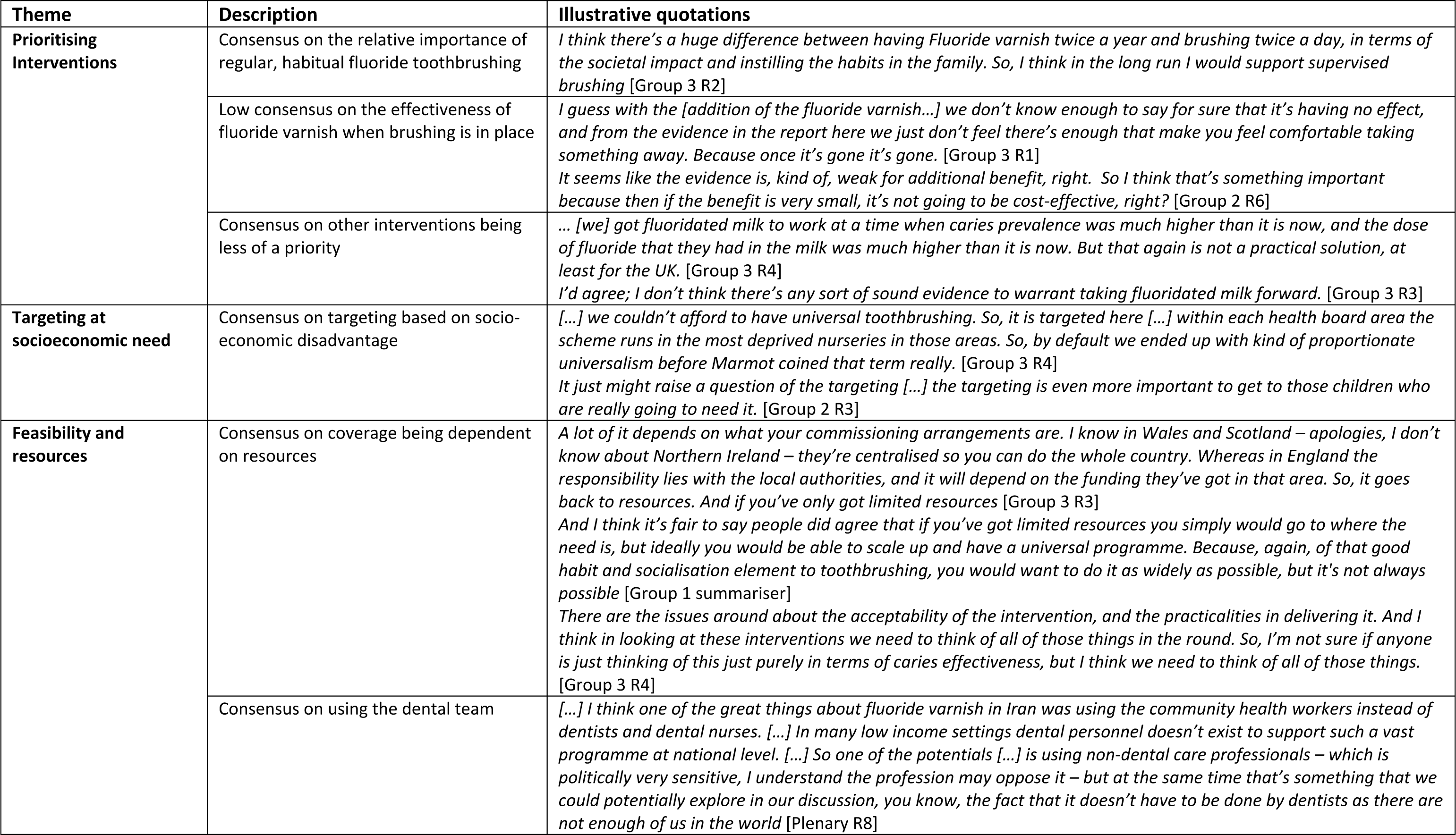
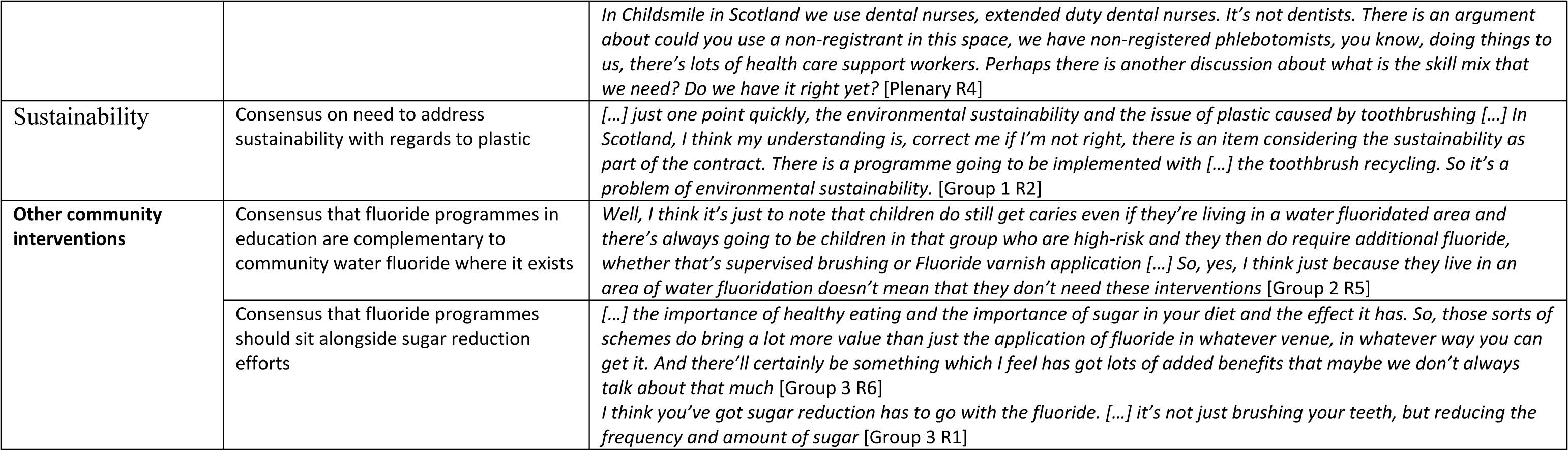
Delphi workshop themes with illustrative quotations.

### Prioritising the intervention

Thematically, there was consensus on supervised toothbrushing with fluoridated toothpaste as the ‘front line’ intervention in the early education setting. As well as being effective and safe, the intervention establishes healthy behaviour from an early age (it was felt studies fail to capture all benefits of supervised brushing e.g. the potential lifelong benefit of forming a healthy habit). This intervention was generally though feasible, but there may be some issues in countries that do not provide free nursery care.

During the workshop, some people felt that the evidence on fluoride varnish, while called into question, hadn’t changed sufficiently to withdraw existing programmes. It was also felt that clinical trials in high-income countries may not show effects that programmes in low-income countries could achieve: “[…] *I’m starting to have real doubts about caries clinical trials in the UK and their ability to show effectiveness. We’ve had a number of big trials now that are showing non-effectiveness”* [Group 3 R4].

### Targeting based on socioeconomic need

The universal approach is supported but experts agree that targeting the most socioeconomically deprived in the first instance should be the priority. With limited resources, experts would prefer to target the most socioeconomically deprived children for longer (to older age) rather than aim for universality (it can be challenging to target schools based on geographical indicators of socioeconomic need).

### Feasibility and resources

Many barriers in implementation were agreed to be important. Whilst toothbrushing might be undertaken supervised without the need for running water (‘dry toothbrushing’), it does require engagement and involvement of teachers or community health workers. Barriers in resources and feasibility terms included: fluoride gels being messy and impractical; the need for refrigeration for fluoridated milk etc. As the group one summariser said: “*it was a very sort of pragmatic discussion. And so rather than a theoretical or esoteric academic discussion, we were talking about things like funding and resources and commissioning.”*

### Sustainability

It was agreed that fluoride interventions in education settings are not currently the most environmentally sustainable programmes. Recycling plastic and examining elements such as staff travel is likely to be a focus in future. However, with regard to environmental sustainability, it was stressed that preventing a significant disease is ultimately the most sustainable thing to do. Efforts to reduce a carbon footprint need to measure the end product such as reduced treatment costs, rather than simply look at the input.

### Other community interventions

The discussions highlighted the importance of a common risk factor approach and how focusing solely on fluoride risks missing benefits of sugar reduction, in particular for wider public health. In areas with water fluoridation, evidence of additional benefit from toothbrushing is emerging. The panel was clear that children still get dental caries in water fluoridated areas and therefore require other interventions.

## Discussion

This project used evidence collected from a systematic overview and Delphi exercise to develop an international expert consensus on fluoride-based interventions in early years educational settings for reducing ECC in young children. We excluded studies of minimally invasive techniques such as Silver Diamine Fluoride (SDF) and Atraumatic Restorative Treatment (ART) from this review because they are curative rather than preventive methods for dental caries. The review of moderate and high-quality papers revealed that supervised toothbrushing delivered in nurseries or primary schools using fluoridated toothpaste containing 1000 to 1450 ppm fluoride, significantly reduced caries and was of high benefit to most disadvantaged children. Panel consensus was that this is still safe and effective in water fluoridated areas and is largely feasible in low/middle-income countries. Fluoride varnish application was generally protective against caries, however, evidence for delivery in an education setting was less clear, especially when delivered alongside supervised toothbrushing.

The evidence for all other modalities of fluoride interventions (fluoride rinse, gel and foam, fluoride supplements, fluoridated milk and fluoridated salt) in nurseries/kindergartens or primary/elementary schools was not supported with high-quality evidence and the information on adverse effects of such interventions was limited.

Results supported the recommended actions set by the WHO Oral Health Draft Global Oral Health Action Plan (2023–2030), i.e. actions 27 to 31, which highlight the importance of delivering toothbrushing programmes to reduce dental caries prevalence and using school settings as a platform for securing broader access to children at early ages [40].

The Delphi exercise allowed for these results to be fed into the survey and discussions with international experts. The literature broadly matched their existing opinions on effective and feasible interventions. The panel agreed toothbrushing with fluoridated toothpaste should be universal for all children in early years education which is supported by WHO recommended actions for the coming decades. [40] [2] The panel was in agreement that toothbrushing is a good vehicle for promoting health behaviour and that, for example, sugar restriction interventions beyond fluoride were important but evidence for restriction per se is limited. [41] The support for supervised brushing with fluoridated toothpaste held even with respect to areas where there is optimal water fluoridation. [42]

Resource issues were felt to be serious, including demands on staff and curricular time. Additionally, political support and priorities, and monitoring of outcomes were felt to be important. Lack of political priority and oral health data are key global challenges to Universal Oral Health Coverage [43].

The experts also raised issues related to education partners (headteachers and class teachers prioritising curriculum activities, availability of school facilities, and fear of cross-infection post-COVID-19), and the importance of solving plastic recycling issues to attain better long-term environmental sustainability, which also required political and governmental support to be resolved. Sustainability and recycling of plastic is becoming more of an important consideration and should be considered in the light of long-term benefits.

### Conclusion

In conclusion, daily supervised toothbrushing with fluoride toothpaste emerged to be the highest priority intervention for preventing early childhood dental caries in early education (nurseries/kindergarten and school) settings. Ideally, this intervention would be universal and extended to older children, but resource, sustainability and feasibility issues are important considerations. Providers need to consider whether there is sufficient added benefit (in terms of effectiveness and cost-effectiveness) from fluoride varnish programmes in education settings where supervised toothbrushing with fluoridated toothpaste is in place.

## Data Availability

All relevant data are within the manuscript and its Supporting Information files.

## Acknowledgement

The authors would like to acknowledge:

The Borrow Foundation for having provided valuable funds for the project.

Childsmile Programme staff for additional support, funded by the Scottish Government Health Directorate.

Emma Pacey, University of Cambridge Department of Health and Social Care for significant help and contribution to data screening.

## Supporting Information

**S1 Table.** Studies excluded from the systematic overview at full text stage

**S2 Table.** Data extraction and quality appraisal for trials and observational studies (n= 61)

**S3 Table.** Data extraction and quality appraisal for reviews (n= 13)

